# Using Primary Care Text Data and Natural Language Processing to Monitor COVID-19 in Toronto, Canada

**DOI:** 10.1101/2022.04.27.22274400

**Authors:** Christopher Meaney, Rahim Moineddin, Sumeet Kalia, Babak Aliarzadeh, Michelle Greiver

## Abstract

**Objective:** To investigate whether a rule-based natural language processing (NLP) system, applied to primary care clinical text data, can be used to monitor COVID-19 viral activity in Toronto, Canada.

**Design:** We employ a retrospective cohort design. We include primary care patients with a clinical encounter between January 1, 2020 and December 31, 2020 at one of 44 participating clinical sites.

**Setting and Context:** The study setting is Toronto, Canada. During the study timeframe the city experienced a first wave of COVID-19 in spring 2020; followed by a second viral resurgence beginning in the fall of 2020.

**Methods and Data:** Study objectives are descriptive. We use an expert derived dictionary, pattern matching tools and a contextual analyzer to classify documents as 1) COVID-19 positive, 2) COVID-19 negative, or 3) unknown COVID-19 status. We apply the COVID-19 biosurveillance system across three primary care electronic medical record text streams: 1) lab text, 2) health condition diagnosis text and 3) clinical notes. We enumerate COVID-19 entities in the clinical text and estimate the proportion of patients with a positive COVID-19 record. We construct a primary care COVID-19 NLP-derived time series and investigate its correlation with other external public health series: 1) lab confirmed COVID-19 cases, 2) COVID-19 hospitalizations, 3) COVID-19 ICU admissions, and 4) COVID-19 intubations.

**Results:** Over the study timeframe 1,976 COVID-19 positive documents, and 277 unique COVID-19 entities were identified in the lab text. 539 COVID-19 positive documents and 121 unique COVID-19 entities were identified in the health condition diagnosis text. And 4,018 COVID-19 positive documents, and 644 unique COVID-19 entities were identified in the clinical notes. A total of 196,440 unique patients were observed over the study timeframe, of which 4,580 (2.3%) had at least one positive COVID-19 document in their primary care electronic medical record. We constructed an NLP-derived COVID-19 time series describing the temporal dynamics of COVID-19 positivity status over the study timeframe. The NLP derived series correlates strongly with external public health series under investigation.

**Conclusions:** Using a rule-based NLP system we identified hundreds of unique COVID-19 entities, and thousands of COVID-19 positive documents, across millions of clinical text documents. Future work should continue to investigate how high quality, low-cost, passively collected primary care electronic medical record clinical text data can be used for COVID-19 monitoring and surveillance.

## 1 Introduction

The coronavirus disease 2019 (COVID-19) pandemic is caused by the viral respiratory pathogen severe acute respiratory syndrome coronavirus-2 (SARS-CoV2). Globally it is estimated that over 500 million SARS-CoV2 infections have occurred and over that 6.2 million people have died because of COVID-19 [World Health Organization: URL accessed April 27, 2022 https://covid19.who.int/]. In many countries COVID-19 remains a persistent threat to human health/well-being, with the capability of inducing continued human, social, and economic loss.

Currently, a confluence of viral, socio-behavioural, and economic factors create a landscape which potentially exacerbates viral transmission and places populations at continued risk of COVID-19 infection. Outside of human control, there exists considerable uncertainty regarding the natural evolution of the SARS-CoV2 virus, and novel variants of concern continue to emerge [Darby and Hiscox, 2021, Lauring and Malani, 2021, Walensky et al., 2021]. When dealing with novel COVID-19 variants of concern, the science regarding transmission characteristics, protection conferred from available vaccines or past infection, and clinical presentation/severity resulting from infection are necessarily incomplete/evolving. From a sociological perspective, vaccine hesitancy, misinformation regarding risks/benefits of vaccines, and the “anti-vax” movement threaten efforts to achieve herd immunity against COVID-19 [Callaghan et al., 2021, Forman et al., 2021, Machingaidze and Wiysonge, 2021]. Further, global inequities create a situation where many nations lack the economic resources necessary to procure and distribute vaccines for their populations [Padma et al., 2021]. As the pandemic extends, there is a risk that continued public adherence with non-pharmaceutical interventions aimed at mitigating viral transmission will wane (e.g. mask wearing, social distancing, and “lockdowns”) [Crane et al., 2021, Doogan et al., 2020]. As a result of these aforementioned factors, and a multitude of others, in many regions across the globe COVID-19 may remain a continued threat to community health.

Working from a hypothesis that we will continue to live with the threat of COVID-19 for the foreseeable future, it becomes paramount that governments and public health authorities develop technologies to rapidly identify and respond to viral re-emergence in their local communities. Tools should assist decision makers determine when/how to promote and re-implement non-pharmaceutical interventions, when to increase procurement/distribution of vaccine inventories and other COVID-19 anti-viral medications, and when/how to expand hospital/ED/ICU capacity. Throughout the pandemic public health researchers have sought novel scientific technologies/data-sources which may act as leading indicators of viral activity, including: cell phone mobility data [Buckee et al., 2020], search history and social media posts [Lampos et al., 2021], data from contact tracing application [Budd et al., 2020], environmental data sources (e.g. COVID-19 RNA from wastewater sources) [Peccia et al., 2020], and health system electronic medical record data [Chapman et al., 2020, Liu et al., 2021].

In this study we seek to investigate whether expressive clinical narrative data, passively captured from primary care patient electronic medical records, can be used as an indicator of COVID-19 viral activity in Toronto, Canada. We apply a rule-based COVID-19 natural language processing (NLP) system to infer the COVID-19 positivity status of primary care text records. Preliminary study objectives relate to the identification of COVID-19 entities in clinical text streams, assessment of COVID-19 positivity status at a document/patient level, and exploration of the agreement in COVID-19 positivity status across mined primary care text streams. The primary objective of the study involves the construction of a primary care NLP-derived COVID-19 indicator series, and assessment of its correlation with other available COVID-19 series (obtained from Toronto Public Health), including: 1) lab confirmed COVID-19 cases, 2) COVID-19 hospitalizations, 3) COVID-19 ICU admissions, and 4) COVID-19 intubations.

## 2 Methods

### 2.1 Study Setting and Context

The study setting is Toronto, Canada. Toronto is the fourth largest metropolitan city in North America and is one the most multicultural and socio-economically diverse cities in the world. Since the WHO declared COVID-19 a pandemic of global concern in March 2020, Toronto has undergone six distinct waves of COVID-19 infection and at times the city has been considered a regional COVID-19 “hotspot”. As of writing (April 2022) Toronto has recorded >318k lab confirmed COVID-19 infections and recorded >4.2k deaths https://www.toronto.ca/home/covid-19/covid-19-pandemic-data/covid-19-weekday-status-of-cases-data/. Provincial/municipal governments have attempted to mitigate pandemic related threats via a combination of non-pharmaceutical interventions, including: promotion of basic infection control measures (e.g. hand washing, use of face masks and other personal protective equipment), contact tracing, social distancing and when necessary strict lockdown measures (i.e. school/business closures, restrictions on large-scale gatherings, and inter-jurisdictional travel advisories). Beginning in 2021, Toronto initiated a mass vaccination campaign, and as of writing (April 2022) Toronto has one of the highest vaccination uptake rates in the world (>91.7% of residents aged 12+ with 1 vaccine dose, >89.2% residents aged 12+ with 2 vaccine doses, and >56.7% of residents aged 12+ with 3 vaccine doses) https://www.toronto.ca/home/covid-19/covid-19-pandemic-data/covid-19-vaccine-data/. Currently, there exists considerable uncertainty regarding novel COVID-19 variants of concern, long-term vaccine effectiveness (including the need for continued booster shots and/or herd immunity), and the possibility of additional waves of COVID-19 infection in 2022 (and beyond).

### 2.2 Study Design, Data Sources, and Inclusion/Exclusion Criteria

The study employs a retrospective open cohort design. The study start date is January 1, 2020 and the study end date is December 31, 2020. Data are collected from primary care physicians (and their associated patients) contributing data to the University of Toronto Practice Based Research Network (UTOPIAN: https://dfcm.utoronto.ca/utopian). Data are collected at a record/ encounter level, with (potentially multiple) encounters hierarchically nested within patients, physicians, and clinics.

Several record identification variables (ID variables) are used in the study, including encounter-level IDs (bill ID, lab ID, health condition diagnosis ID, and clinical note ID), patient ID, physician ID and site/clinic ID. Three main text data streams are utilized in the study: 1) lab text, 2) health condition diagnosis text, and 3) clinical notes. Each of the text data streams are measured on an encounter level. The lab text stream contains narrative information associated with laboratory test orders/results. The health condition diagnosis text stream contains diagnoses and other important clinical events that can be considered as past medical history for the patient. The clinical notes text stream contains narrative data from clinical visits/encounters, referrals, consults and reports. In addition to the clinical text data used in this study, patient age and sex are two demographic variables used for characterizing the cohort under investigation.

We include all records (and associated patients) who experience at least one clinical encounter with the primary healthcare system over our study timeframe (i.e. January 1, 2020 through December 31, 2020). We define a valid encounter as an observed billing ID, lab ID, health condition diagnosis ID, or clinical note ID occurring over the study period which generates a non-null data value. Using site/clinic level postal information we include only sites from Toronto, Canada (leading character M in the postal code). We exclude patients if they are missing age, sex, or postal code.

### 2.3 A Natural Language Processing System for COVID-19 Document Classification and Biosurveillance

We utilize a rule based natural language processing (NLP) system to identify (at an encounter/document level) primary care text streams containing utterances pertaining to COVID-19 [Chapman et al., 2020]. The NLP system uses pattern matching and expert curated dictionaries/gazetteers to identify COVID-19 related entities in clinical text streams. In addition, the system employs the ConText algorithm [Harkema et al., 2009] to disambiguate the linguistic circumstances under which the COVID-19 entity occurred (e.g. who experienced the COVID-19 related entity, whether the COVID-19 related entity occurred or was negated, whether the utterance was not related to a patient COVID-19 clinical diagnosis, and generally, whether there existed uncertainty regarding the COVID-19 entities occurrence). At a document level, each primary care clinical record is classified as: 1) COVID-19 positive, 2) COVID-19 negative, or 3) unknown COVID-19 status. We apply the COVID-19 biosurveillance tool to three clinical text streams from the UTOPIAN primary care electronic medical record system: 1) lab text, 2) health condition diagnosis text, and 3) clinical notes.

The tool was originally developed and validated at the United States Veteran’s Affairs health system [Chapman et al., 2020]. The operating characteristics of the NLP system were exceptional, with internal validation reporting 94.2% sensitivity and 82.4% positive predictive value. We conducted a small validation study on each of the UTOPIAN text data streams to further investigate sensitivity, specificity, positive predictive value and negative predictive value in our local setting. The results of this small validation study are given in Appendix A. Further details regarding computational implementation of the COVID-19 biosurveillance tool are described at the authors GitHub repository https://github.com/abchapman93/VA_COVID-19_NLP_BSV.

### 2.4 Statistical Methods

We identify COVID-19 entities in each primary care clinical text stream. At a record level we classify each free text clinical document as 1) COVID-19 positive, 2) COVID-19 negative, or 3) unknown COVID-19 status. We report descriptive characteristics regarding the number/proportion of positive COVID-19 documents at an encounter-level. We aggregate encounter level COVID-19 document classifications to a patient-level and describe the number of patients whose primary care clinical notes contain at least one positive COVID-19 utterance. At a patient-level, we cross-tabulate derived COVID-19 positive indicator vectors measured from each of the text streams and describe agreement in COVID-19 ascertainment across data sources. Finally, we investigate (at a patient-level) whether demographic variables such as age, sex, or clinic location are associated with COVID-19 positivity status.

Using descriptive time series methods, we investigate temporal variation in the number of COVID-19 positive documents in the electronic medical record over our study time frame. We discretize time in 53 weekly bins, from January 1, 2020 through December 31, 2020. We create an overall count of the number of COVID-19 positive utterances/occurrences by summing the number of positive document classifications from each of the three text streams (1. lab text, 2. health condition diagnosis text, and 3. clinical notes). Using descriptive time series plots we visually compare how our NLP derived primary care COVID-19 positivity series correlates with other known COVID-19 time series externally derived from Toronto Public Health, including: 1) lab confirmed COVID-19 cases, 2) COVID-19 hospitalizations, 3) COVID-19 ICU admissions, and 4) COVID-19 intubations [Toronto Public Health, 2022].

### 2.5 Ethics

This study received ethics approval from North York General Hospital Research Ethics Board (REB ID: NYGH 20-0014).

## 3 Results

### 3.1 Descriptive Characteristics of Study Sample

The study timeframe is January 1, 2020 through December 31, 2020. We include data from 44 primary care clinics in Toronto, Canada. During the study timeframe 196,440 patients had contact - in-person, email, phone or video - with their primary care provider (resulting in a recorded billing ID, lab ID, health condition diagnosis ID or clinical note ID in the patient primary care electronic medical record). The majority of observed patients observed in the sample are female (58.8%). In terms of age structure: 14.1% of patients are 0-18 years old, 27.1% of patients are 18-40 years old, 37.2% of patients are 40-65 years old, 18.4% of patients are 65-85 years old and 3.2% of patients are >85 years old. Table 1 describes demographic characteristics associated with lab text, health condition diagnosis text and clinical notes.

**Table 1:** Descriptive characteristics of lab texts, health condition diagnosis texts and clinical notes included in the study sample, measured on a record/encounter-level.

|  | Lab Text<br>(N=2,360,957 records) | Health Condition Text<br>(N=107,075 records) | Clinical Notes<br>(N=454,503 records) |
| --- | --- | --- | --- |
| Age |  |  |  |
| 0-18 years | 40,522 (1.7%) | 7,800 (7.3%) | 34,324 (7.6%) |
| 18-40 years | 420,851 (17.8%) | 25,887 (24.2%) | 101,081 (22.2%) |
| 40-65 years | 997,457 (42.3%) | 41,110 (38.4%) | 170,701 (37.6%) |
| 65-85 years | 770,285 (32.6%) | 27,398 (25.6%) | 120,888 (26.6%) |
| >85 years | 131,842 (5.6%) | 4,880 (4.5%) | 27,509 (6.0%) |
| Sex |  |  |  |
| Male | 948,508 (40.2%) | 40,040 (37.4%) | 164,780 (36.2%) |
| Female | 1,412,449 (59.8%) | 67,035 (62.6%) | 289,723 (63.8%) |

### 3.2 COVID-19 Clinical Entities and Document Classifications in Primary Care Clinical Text Streams

We identify a diverse variety of COVID-19 entities in the three primary care electronic medical record text streams investigated in this study. Both the list of specific entities/tokens and the most-frequently occurring entities/tokens identified vary according to the text stream under consideration. Using the ConText algorithm, the COVID-19 biosurveillance system classifies documents as 1) COVID-19 positive, 2) COVID-19 negative, or 3) unknown COVID-19 status. We report the percentage of documents in each COVID-19 category in Table 2 below.

**Table 2:** COVID-19 document classifications and COVID-19 entities/tokens identified from each primary care clinical text stream: lab text, health condition diagnosis text and clinical notes.

|  | Lab Text<br>(N=2,360,957 records) | Health Condition Text<br>(N=107,075 records) | Clinical Notes<br>(N=454,503 records) |
| --- | --- | --- | --- |
| COVID-19 Document Classification |  |  |  |
| COVID-19 Positive | 1,976 (0.1%) | 539 (0.5%) | 4,018 (0.9%) |
| COVID-19 Negative | 2,296,462 (97.3%) | 105,380 (98.4%) | 411,093 (90.4%) |
| Unknown COVID-19 Status | 62,519 (2.6%) | 1,156 (1.1%) | 39,392 (8.7%) |
| Number of Unique COVID-19 Entities | 277 | 121 | 644 |
| Most Frequent COVID-19 Entities |  |  |  |
| Rank 1 | COVID-19 | COVID-19 | COVID |
| Rank 2 | COVID19 | COVID | covid |
| Rank 3 | Novel Coronavirus | COVID-19 Confirmed | COVID-19 |
| Rank 4 | CoronaVirusN2019 | covid | Covid |
| Rank 5 | COVID | COVID19 | Covid-19 |
| Rank 6 | Novel coronavirus | Covid-19 | COVID19 |
| Rank 7 | SARS-CoV-2 | Covid | covid-19 |
| Rank 8 | 2019-nCoV | COVID positive | coronavirus |
| Rank 9 | coronavirus | COVID+ | COVID-19 coronavirus |
| Rank 10 | 2019-nCov | Covid +ve | covid19 |

### 3.3 Patient Level COVID-19 Status, Agreement of COVID-19 Status by Source, and Correlates of COVID-19

196,440 unique patients had at least one clinical event recorded in their primary care electronic medical record over our study timeframe (i.e. January 1, 2020 through December 31, 2020). A total of 1,573 (0.8%) of these patients had a COVID-19 positive lab text in their medical record, 516 (0.3%) of these patients had a COVID-19 positive health condition diagnosis text in their medical record, and 3,022 (1.5%) had a COVID-19 positive clinical note in their medical record. We constructed a binary composite COVID-19 status variable (at a patient-level), based on whether any of the text streams under consideration indicate COVID-19 positive status. A total of 4,580 (2.3%) patients with at least one primary care contact/record during the initial pandemic year (January 1, 2020 through December 31, 2020) experienced at least one COVID-19 positive document in their electronic medical record.

In this study, we observe increasing COVID-19 positivity status with increasing age. We observe a positive indication of COVID-19 in 1.3% of patient 0-18 years old, 2.4% of patients 18-40 years old, 2.4% of patients 40-65 year old, 2.6% of patients 65-85 years old and 3.3% of patients 85 years or older. In our sample, we observe that females were more likely than males to record a COVID-19 positive diagnosis over our study timeframe (2.5% in females vs. 2.1% in males). Data were collected from 44 clinical sites across Toronto. COVID-19 positivity status varied across geographic regions of Toronto. In one clinic 8.9% of patients observed over the study timeframe had a COVID-19 positive indicator in their electronic medical record; whereas, in the least impacted clinic only 0.09% of patients presenting over the study timeframe had a recorded COVID-19 positive indicator in their electronic medical record.

At a patient level we descriptively evaluate agreement between COVID-19 positive status as estimated across each of the three different electronic medical record text streams: 1) lab text, 2) health condition diagnosis text and 3) clinical notes. Table 3 highlights limited agreement in COVID-19 positivity status across each of the three text streams.

**Table 3:**
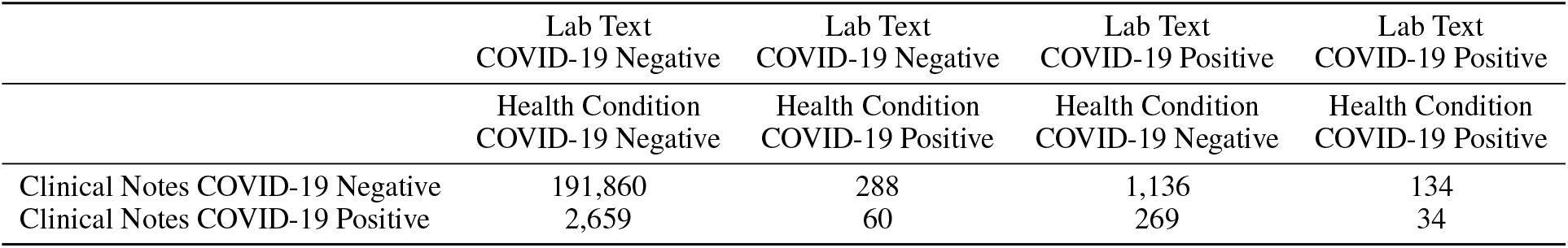
Three-way cross-tabulation comparing the number of COVID-19 positive indications (at a patient-level) from the lab text, health condition diagnosis text and clinical note data streams.

### 3.4 A COVID-19 Primary Care NLP-Derived Time Series Analysis and External Validation

To investigate the temporal dynamics of COVID-19 in primary care electronic medical records over the study period we discretize time into 53 weekly bins and sum the number of COVID-19 positive indications/utterances in each week (from either the lab text, health condition diagnosis text or clinical notes). We visualize the resulting primary care NLP-derived series using several time series plots. We further compare the primary care NLP-derived series against other externally generated COVID-19 time series collected/curated by Toronto Public Health. We descriptively assess the correlation between our NLP-derived primary care COVID-19 series, and four public health series: 1) lab confirmed COVID-19 cases, 2) COVID-19 hospitalizations, 3) COVID-19 ICU admissions, and 4) COVID-19 intubations. While the primary care derived series appear strongly correlated with each of the Toronto Public Health COVID-19 series (using naïve Pearson correlation); in many cases, the series track each other strongly, with one series leading/lagging the other during different time periods of the 2020 pandemic year.

**Figure 1:**
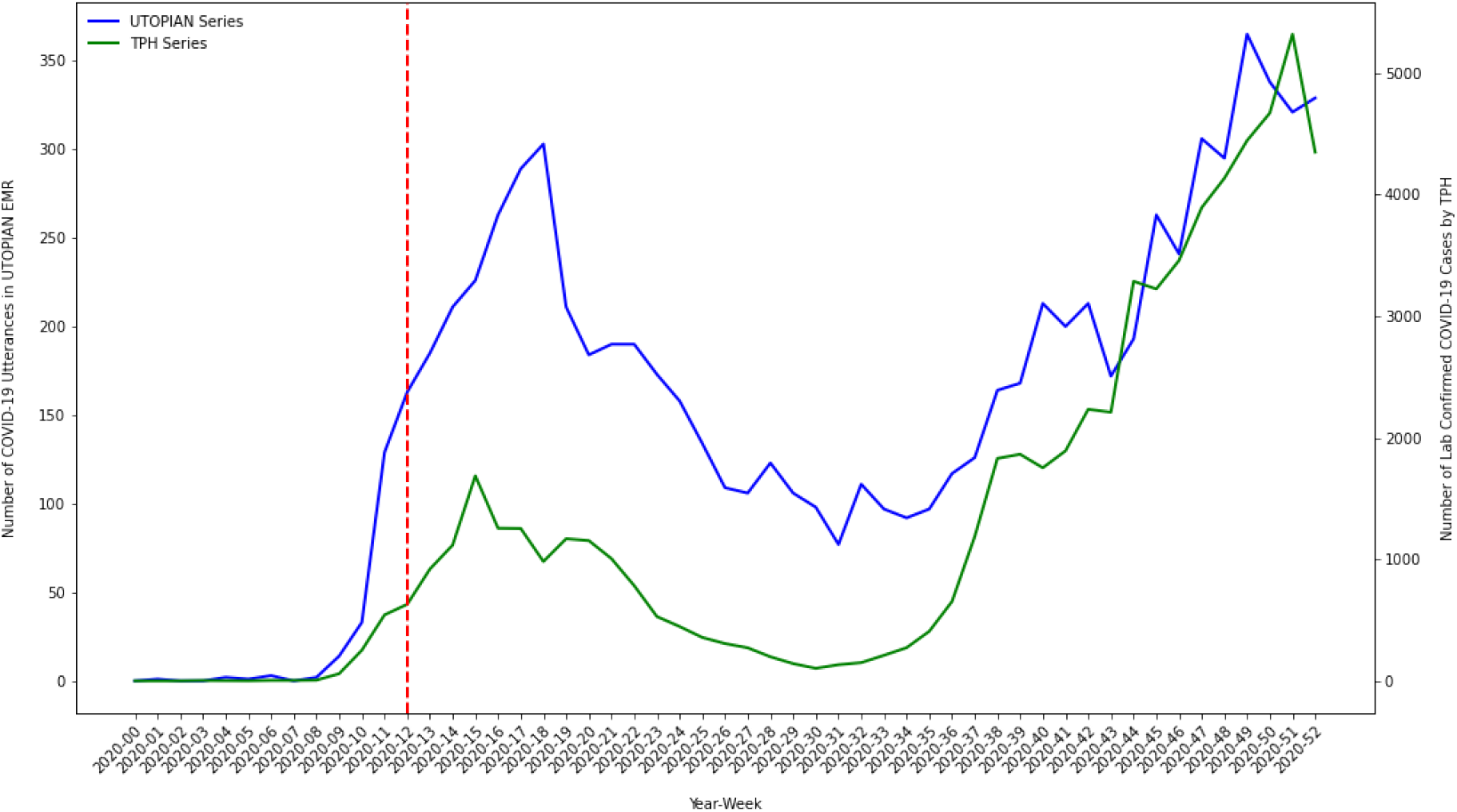
Time series plot of the composite count of the number of COVID-19 positive utterances from each of the three electronic medical records text streams (lab text, health condition diagnosis text and clinical notes) versus the number of lab confirmed COVID-19 cases in Toronto, Canada (data extracted from Toronto Public Health).

**Figure 2:**
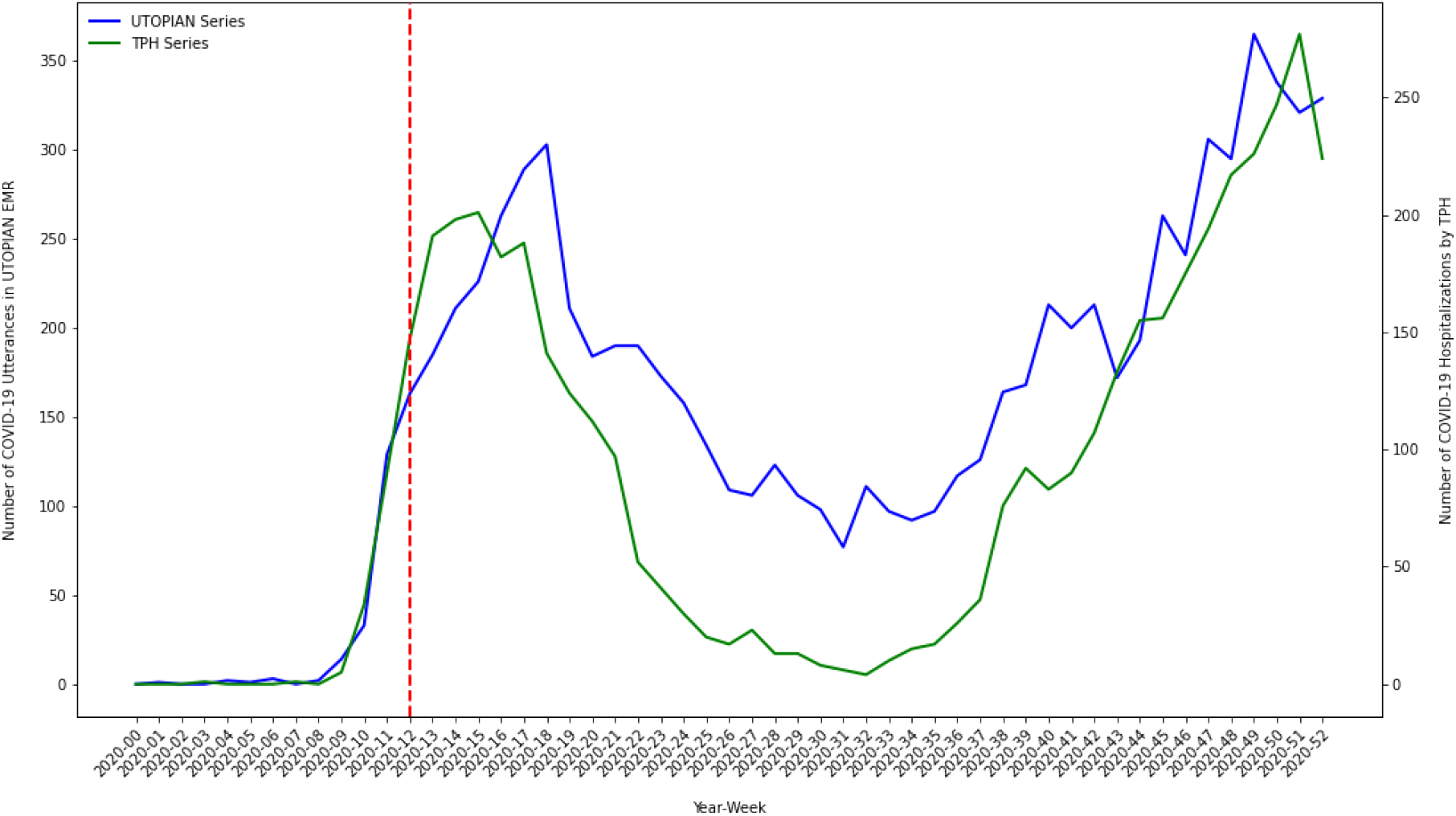
Time series plot of the composite count of the number of COVID-19 positive utterances from each of the three electronic medical records text streams (lab text, health condition diagnosis text and clinical notes) versus the number of COVID-19 related hospital admissions in Toronto, Canada (data extracted from Toronto Public Health).

**Figure 3:**
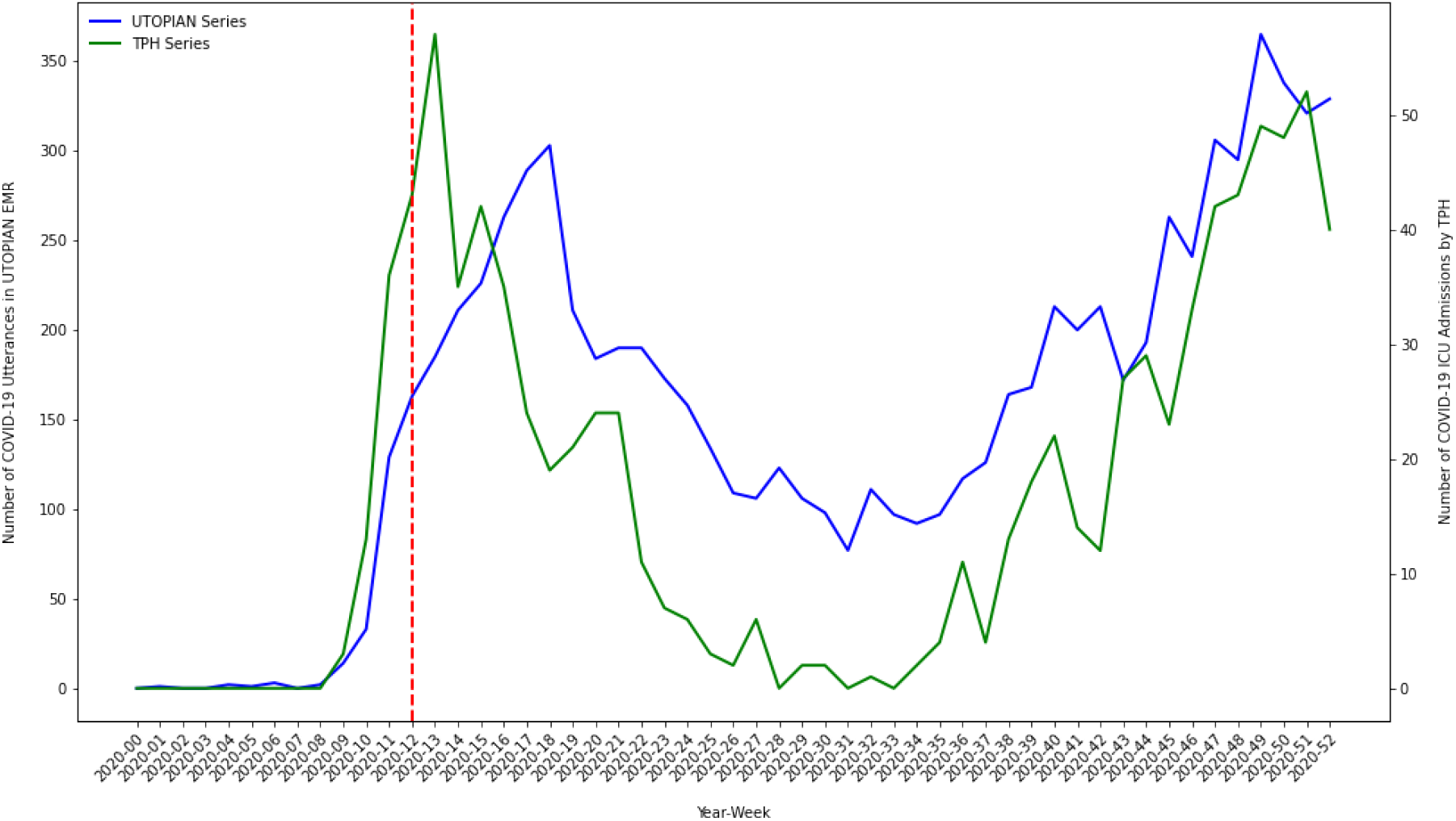
Time series plot of the composite count of the number of COVID-19 positive utterances from each of the three electronic medical records text streams (lab text, health condition diagnosis text and clinical notes) versus the number of COVID-19 related ICU admissions (data extracted from Toronto Public Health).

**Figure 4:**
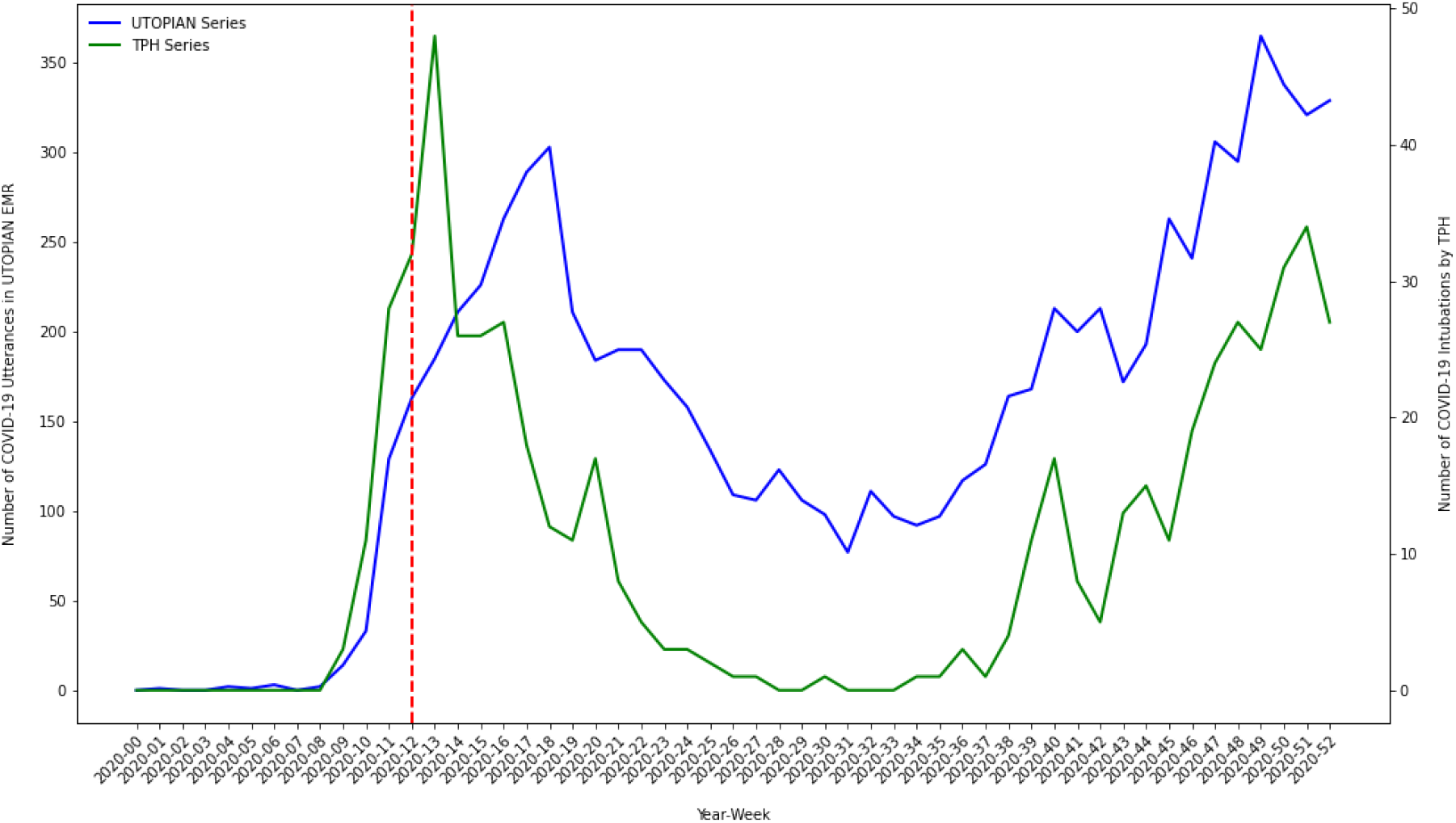
Time series plot of the composite count of the number of COVID-19 positive utterances from each of the three electronic medical records text streams (lab text, health condition diagnosis text and clinical notes) versus the number of COVID-19 related intubations in Toronto, Canada (data extracted from Toronto Public Health).

## 4 Discussion

In this study we applied a rule-based NLP system to identify COVID-19 positive utterances over millions of clinical text documents, gathered from hundreds of thousands of primary care patient records. The NLP system was originally developed by the United States Veteran’s Affairs heath system, and we transported the technology to primary care electronic medical records from Toronto, Canada. The rule-based NLP system identified hundreds of distinct COVID-19 entities across three diverse clinical text streams: lab text, health condition diagnosis text and clinical notes. Thousands of primary care patients were identified using the NLP system as having at least one positive COVID-19 document classification over our study timeframe (i.e. January 1, 2020 through December 31, 2020).

The study findings corroborate the important role that primary care physicians have played during the pandemic. Buried within the primary care electronic medical record, there exist thousands of instances where COVID-19 related entities are mentioned in the context of medical communication, disease prevention, disease management and care-planning. A key finding relates to the diversity of COVID-19 entities discovered and the heterogeneity in which these entities are recorded in modern electronic medical record systems. Study findings suggest that COVID-19 utterances are being recorded in numerous areas of the primary care electronic medical record system where clinical narrative data are captured, including: lab text, health condition diagnosis text and clinical notes. Further, there does not appear to be strong agreement between patient-level COVID-19 positivity status across these clinical text data sources. This finding suggests that COVID-19 related information is not captured/recorded in a single primary care electronic medical record field, and that studies employing primary care electronic medical records to identify COVID-19 positivity status would be wise to mine all available data sources/streams available, as reliance on a single source is likely to miss important COVID-19 utterances mentioned elsewhere in the patient medical record.

A primary objective of this study was to investigate the temporal dynamics of inferred COVID-19 positivity status from primary care electronic medical records. The estimated COVID-19 primary care NLP-derived time series exhibit temporal dynamics which have strong face validity over the study period. Early in 2020 (prior to the WHO declaring COVID-19 a global pandemic) COVID-19 lab confirmed infections and hospitalizations were low (near zero) in Toronto – this is corroborated in our analyses. In March/April 2020 we observed a sharp increase in COVID-19 positive documents identified in the primary care electronic medical record – the timing being consistent with the first wave of COVID-19 infection in Toronto, Canada. COVID-19 positive document counts displayed a low/minima in the summer of 2020 (as Toronto moved out of pandemic wave-01, following weeks of governmental imposed lockdowns). As lockdown measures were relaxed and public adherence to non-pharmaceutical interventions waned through the fall of 2020, we observed a sharp increase in the number of COVID-19 positive documents/patients recorded in the latter part of 2020 (which is consistent with the commencement of the wave-02 COVID-19 infection surge in Toronto). We plotted our COVID-19 NLP derived series against other important COVID-19 indicator series independently generated by Toronto Public Health using public-health/hospital data sources, including: 1) lab confirmed COVID-19 cases, 2) COVID-19 hospitalizations, 3) COVID-19 ICU admissions, and 4) COVID-19 intubations. On external validation, our primary care NLP-derived COVID-19 series correlated strongly with other important public health series. Strong empirical correlations are encouraging, and future work should continue to investigate whether primary care electronic medical record data can be used in the quest to uncover leading indicators of COVID-19 infection/hospitalization – as this would greatly assist public health officials and hospital executives attempting to plan for future waves of COVID-19 infection.

A unique aspect of our study is the use of free text primary care electronic medical record data to investigate COVID-19 positivity status, and temporal dynamics of COVID-19 over the initial pandemic year. Few other studies/jurisdictions have utilized primary care electronic medical record data for COVID-19 phenotyping or for the identification of leading indicators of COVID-19 infection/hospitalization. The work by Chapman et al. [2020] provides a working tool for performing COVID-19 biosurveillance using hospital electronic medical record data. Liu et al. [2021] developed a COVID-19 “hot-spot” identification algorithm based on the presence/absence of 10 clinical indicators, in the context of a large integrated health system in California, USA. The authors evaluated their algorithm using similar design/methods as our study and found their method to be a strong leading indicator of COVID-19 cases/hospital-admissions. The ability to reconstruct their bespoke algorithm in our primary care setting would be challenging and would require a combination of remapping clinical disease codes to our nomenclature and the use of NLP named entity recognition tools to extract clinical indicators not collected using our standardized nomenclatures. Many other studies have investigated COVID-19 phenotyping using electronic medical record data. Certain exceptional studies include Brat et al. [2020] and Klann et al. [2021]; however, these studies were developed in the context of hospital electronic medical records, and the availability of requisite data sources for phenotype construction in primary care settings may be limited.

Our study is not without limitations. A chief limitation of our study is that we employ a rule based COVID-19 phenotype algorithm which has undergone minimal internal validation in our local setting (see Appendix A). A challenge with internal validation of COVID-19 phenotyping algorithms relates to sampling design. As illustrated in Appendix A, evaluation of algorithm operating characteristics based on a random sample of clinical notes results in the identification of only a small number of true positive COVID-19 documents (and similarly a small number of algorithm predicted positive documents); hence, internal estimates of sensitivity and positive predictive value are imprecise. Estimation of algorithm operating characteristics from large samples are associated with increased costs (financial costs, time costs and human-resource costs), however, are necessary to achieve precise estimates of sensitivity and positive predictive value. Alternatively, sequential or stratified sampling designs may provide more efficient estimates of algorithm operating characteristics as compared to simple random sampling designs. That said, the original authors did perform their own validation of the COVID-19 biosurveillance tool and report exceptional sensitivity (94.2%) and positive predictive value (82.4%) [Chapman et al., 2020]. Qualitatively, our small validation study suggests similarly effective performance in an entirely different study setting/context. A central objective of our study involved a rigorous external validation of the COVID-19 biosurveillance system against important public-health/hospital-based indicators of COVID-19 activity in Toronto, Canada. Our study findings have face validity, as counts of documents reporting positive utterances of COVID-19 correlate strongly against externally generated COVID-19 indicator series collected from Toronto Public Health. Another limitation of our study relates to the retrospective design and delay/lag in data reception, curation and reporting in our local jurisdiction. The utility of our findings would be enhanced if more timely access to primary care electronic medical record data existed in our jurisdiction, which would facilitate using our biosurveillance tool for near real-time monitoring/surveillance of COVID-19 in Toronto, Canada.

## 5 Conclusions

In this study, we applied a rule-based NLP system for COVID-19 biosurveillance to our primary care electronic medical records in Toronto, Canada. The method scaled to millions of primary care clinical text documents, and hundreds of thousands of patient records. We identified hundreds of unique COVID-19 entities, and thousands of COVID-19 positive clinical documents, across three primary care clinical text streams. The resulting primary care COVID-19 NLP-derived time series correlated strongly against other important COVID-19 indicator series, externally generated from hospital/public-health data sources. Future work should continue to investigate whether leading indicators of COVID-19 cases or hospitalizations may be uncovered using readily available, passively collected, high quality primary care electronic medical record data.

## Data Availability

All data produced in the present study are available upon reasonable request to the authors.

https://dfcm.utoronto.ca/utopian

## Appendix A: UTOPIAN Validation of the USVA COVID-19 Biosurveillance System [Chapman et al., 2020]

We conducted a small validation study to investigate operating characteristics of the COVID-19 biosurveillance system developed by Chapman et al. [2020] in the context of our study setting - primary care clinical practices in Toronto, Canada. Using data from the United States Veteran’s Affairs health system, Chapman et al. [2020] estimate the sensitivity of their COVID-19 biosurveillance algorithm to be 94.2% and the positive predictive value to be 82.4%.

For each of the three text streams under consideration in our study (1. lab text, 2. health condition diagnosis text, and 3. clinical notes) we randomly sample N=500 documents for inclusion in our internal validation study. A single researcher/biostatistician (CM) reviewed each of the randomly sampled free text notes and labelled them as COVID-19 positive versus COVID-19 negative. For each of the three text streams, we compared the human labelled COVID-19 classifications versus the algorithm derived COVID-19 classifications. We report the findings from our small internal validation study in Appendix Tables 4-7.

**Table 4:**
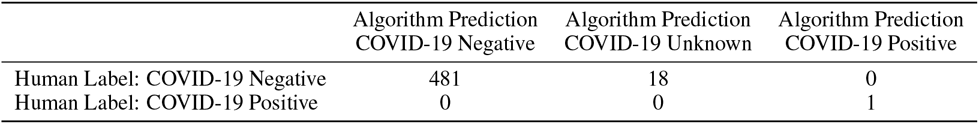
Internal validation of the COVID-19 biosurveillance system relative to the UTOPIAN lab text data source.

For the lab text stream (Table 4), only a single positive COVID-19 note was identified on human review, which was also identified as positive by the COVID-19 biosurveillance algorithm. In the lab text, 18 notes were identified as unknown COVID-19 status by the biosurveillance algorithm, all of which were identified as negative on human review. Several of these 18 notes represented truly negative lab reports, where COVID-19 lab tests identified the virus as being “not detected” on evaluation. Another subset of the unknown notes were COVID-19 lab requisitions for which no positive/negative indication was provided in the text (i.e. labs were ordered and results were pending or undocumented in the text).

For the health condition diagnosis text stream (Table 5), human review identified 8 documents with a (presumed) positive COVID-19 diagnosis; of which only 4 were identified by COVID-19 biosurveillance algorithm as being COVID-19 positive documents. When writing notes in the health conditions diagnosis text stream, physicians tend to be very brief/terse. For example, the 4 document which the COVID-19 biosurveillance algorithm did not identify as being positive (but human review did identify as being positive) all took the format: “COVID-19 [date]” (which we assumed was indicating the date of a positive COVID-19 diagnosis at the note/patient-level). In these cases the contextual analyzer [Harkema et al., 2009] does not have enough information to definitely identify whether a note/patient is COVID-19 positive or not; at which point it defaults to an “unknown” classification status.

**Table 5:** Internal validation of the COVID-19 biosurveillance system relative to the UTOPIAN health condition diagnosis text data source.

|  | Algorithm Prediction<br>COVID-19 Negative | Algorithm Prediction<br>COVID-19 Unknown | Algorithm Prediction<br>COVID-19 Positive |
| --- | --- | --- | --- |
| Human Label: COVID-19 Negative | 490 | 2 | 0 |
| Human Label: COVID-19 Positive | 0 | 4 | 4 |

For the clinical notes text stream (Table 6), human review again identified a single COVID-19 positive document, which was also identified as being COVID-19 positive by the biosurveillance algorithm. For the clinical notes text stream the COVID-19 biosurveillance algorithm also identified 63 documents as unknown COVID-19 status. The majority of these documents contained boilerplate text regarding appointments occurring via phone/video/email as a result of the COVID-19 pandemic. Other documents of unknown COVID-19 status contained utterances regarding COVID-19 which pertained to individuals other than the patient (e.g. wife diagnosed with COVID-19) or for which COVID-19 status was uncertain (e.g. individual was symptomatic for COVID-19 and instructed to report to an assessment center for further evaluation).

**Table 6:**
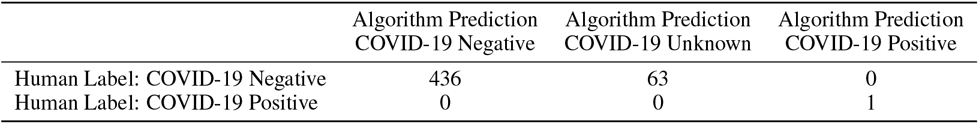
Internal validation of the COVID-19 biosurveillance system relative to the UTOPIAN clinical note data source.

Overall, the COVID-19 biosurveillance algorithm developed by Chapman et al. [2020] performed well in identifying COVID-19 positive documents in our primary care setting, making few false positive or false negative errors. As expected, our internal validation study resulted in too few instances of human-labelled/algorithm-predicted COVID-19 positive documents, under a random sampling design, to generate precise estimates of algorithm operating characteristics. Hence internal estimates of sensitivity and positive predictive value from this study are necessarily imprecise (especially when interpreted at the level of the individual text streams). In Table 7, we aggregate each of the N=500 independent documents, across each of the three text streams (1. lab text, 2. health condition diagnosis text, and 3. clinical notes). We assumed “COVID-19 positive” documents were positive; whereas, “COVID-19 negative” documents and “COVID-19 unknown” documents were combined into a single “non-positive/negative” category. Under this assumption, we compute overall operating characteristics of the COVID-19 biosurveillance algorithm developed by Chapman et al. [2020] in our local study setting. Sensitivity was estimated as 6/10=60.0% (95% exact binomial confidence interval: 26.6% - 87.8%); specificity was estimated as 1490/1490=100.0% (95% exact binomial confidence interval: 99.8% - 100.0%); positive predictive value was estimated as 6/6=100.0% (95% exact binomial confidence interval: 54.1% - 100.0%); and negative predictive value was estimated as 1490/1494=99.7% (95% exact binomial confidence interval: 99.3% - 99.9%).

**Table 7:** Internal validation of the COVID-19 biosurveillance system relative to the aggregated UTOPIAN lab text, health condition diagnosis text and clinical note data sources (N=1500 independent free text documents).

|  | Algorithm Prediction<br>COVID-19 Negative | Algorithm Prediction<br>COVID-19 Unknown | Algorithm Prediction<br>COVID-19 Positive |
| --- | --- | --- | --- |
| Human Label: COVID-19 Negative | 1407 | 83 | 0 |
| Human Label: COVID-19 Positive | 0 | 4 | 6 |

Future work will consider estimating the operating characteristics of the COVID-19 biosurveillance system algorithm in the UTOPIAN primary care study setting using a larger validation sample, such that sensitivity and positive predictive value can be estimated with increased precision. We will also consider validation studies which employ multiple raters to label documents (rather than a single rater - CM). Finally, we will investigate alternative designs for estimating operating characteristics of phenotyping algorithms applied to rare diseases (i.e. disease conditions with low prevalence).

## Notes

### Competing Interest Statement

The authors have declared no competing interest.

### Funding Statement

This study did not receive any funding.

